# Prevention of household transmission crucial to stop the catastrophic spread of COVID-19 in cities

**DOI:** 10.1101/2020.06.05.20123711

**Authors:** Noel Gutiérrez Brizuela, Humberto Gutiérrez Pulido, Kimberlyn Roosa, Néstor García Chan, Jorge Hernández-Bello, José Francisco Muñoz-Valle, Gabriela Macedo-Ojeda, Guillermo González-Estevez, Javier Alonso López-Chávez, Ricardo Villanueva-Lomelí, Gerardo Chowell Puente

**Affiliations:** Scripps Institution of Oceanography, University of California San Diego, La Jolla, California, USA; Centro Universitario de Ciencias Exactas e Ingeniería, Universidad de Guadalajara, Guadalajara, Jalisco, Mexico; School of Public Health, Georgia State University, Atlanta, Georgia, USA; Centro Universitario de Ciencias de la Salud, Universidad de Guadalajara, Guadalajara, Jalisco, Mexico; Rectoría General, Universidad de Guadalajara, Guadalajara, Jalisco, Mexico

## Abstract

After weeks under lockdown, metropolitan areas fighting the spread of COVID-19 aim to balance public health goals with social and economic standards for well-being. Mathematical models of disease transmission seeking to evaluate mitigation strategies must assess the possible impacts of social distancing, economic lockdowns and other measures. However, obscure relations between model parameters and real-world phenomena complicate such analyses. Here, we use a high-resolution metapopulation model of Guadalajara (GDL, Western Mexico) to represent daily mobility patterns driven by economic activities and their relation to epidemic growth. Given the prominence of essential activities in the city’s economy, we find that strategies aiming to mitigate the risk of out-of-home interactions are insufficient to stop the catastrophic spread of COVID-19. Using baseline reproduction numbers *R*_0_ = [2.5, 3.0] in the absence of interventions, our simulations suggest that household transmission alone can make *R_t_* ∼ 1, and is estimated to drive 70 *±*15% of current epidemic growth. This sets an upper bound for the impact of mobility-based interventions, which are unlikely to lower *R_t_* below 1.3 and must be complemented with aggressive campaigns for early case detection and isolation. As laboratory testing and health services become insufficient to meet demand in GDL and most other cities, we propose that cities facilitate guidelines and equipment to help people curb spreading within their own homes. Postponing these actions will increase their economic cost and decrease their potential returns.

**Author summary:** Public health strategies to mitigate the spread of COVID-19 in metropolitan areas have focused on preventing transmission in schools, work sites and other public spaces. Here, we use a demographically- and spatially-explicit model of Guadalajara (GDL, Western Mexico) to represent economic lockdowns and their impact on disease spread. Our findings suggest that viral exposure within households accounts for 70*±*15% of the epidemic’s current growth rate. This highlights the importance of early case detection and isolation as necessary measures to prevent the spread of COVID-19 between strangers and close contacts alike.

## Introduction

Metropolitan areas around the world have implemented containment and mitigation measures against the COVID-19 pandemic. Notably, non-essential activities have been interrupted to limit the number of people exposed to the SARS-CoV-2 virus in schools, retail stores, offices, and other places of interest. However, the economic burden exacted by these measures has overwhelmed societies, leading to massive economic losses that further complicate the fight against the coronavirus pandemic. Carefully validated models of pandemic spread that incorporate realistic population mixing patterns could help restart the economy while limiting the spread of the virus.

Recent studies have emphasized the potential benefits of aggressive testing campaigns aimed to find and isolate symptomatic and asymptomatic carriers [1–5], but these measures require a significant amount of resources, present logistical challenges, and become increasingly unattainable in developing nations as incidence soars across metropolitan areas. Although mathematical models of disease transmission can help assess the potential effects of mitigation strategies, the interpretation and communication of simulation results to the broader public is far from trivial [6,7]. In most models, public health interventions and reactive behavior are represented by time-dependent variables that are related to the epidemic’s reproduction number *R_t_*, defined as the number of secondary cases caused by a single patient who becomes infective at time *t* [8,9]. However, without a realistic integration of city-specific demographics and mixing patterns, mathematical models can only assume the relations between mitigation strategies and epidemic growth [10].

In this letter, we develop and evaluate a metapopulation model that incorporates high-resolution census and economic data to replicate economic lockdowns and their relations to COVID-19 transmission in metropolitan areas. Using the Guadalajara Metropolitan Area (GDL, 2020 population estimated at 4.9 million inhabitants) as an example, we find that mitigation strategies that aim to prevent disease spreading in worksites, schools, and public spaces are unlikely to lower *R_t_* below 1.3. However, we estimate that infections within residential settings can explain more than 55% of epidemic growth during economic lockdowns. This suggests that catastrophic spread of COVID-19 can only be avoided with mixed strategies that aim to prevent disease transmission within households and public spaces. As hospital demand soars throughout the world’s cities and the majority of patients go untested, we recommend that governments strive to isolate all symptomatic patients even without laboratory confirmation. Authorities unable to setup designated areas for case isolation must then provide citizens with equipment and guidelines for effective self-isolation at home.

## Public health context

As of June 2, official data indicate that people in the state of Jalisco (*∼* 60% of whom live in GDL) have received a total of 23,019 PCR tests for SARS-CoV-2 (2.8 tests per 1000 inhabitants, ssj.jalisco.gob.mx/prensa/noticia/9075). With 3,088 confirmed cases to date and 1292 that remain unresolved (13.4% positivity rate), the state of Jalisco is thought to have a disproportionately low incidence despite its social, cultural and economic prominence in Mexico. Although confirmed case counts can produce misleading comparisons between outbreaks in different regions due to differences in testing rates, daily series of COVID-19 deaths comprise 100 deaths in GDL (2.3 deaths per 100,000 people) out of 10,637 deaths nationwide (8.4 deaths per 100,000 people).

The first case of COVID-19 in Jalisco was confirmed on March 11; the next day, non-pharmaceutical measures to curb the spread of COVID-19 began in GDL, when all large public events were cancelled (www.jalisco.gob.mx/es/prensa/noticias/102580). The Jalisco State Government later shut the city’s schools and universities on March 17 and issued shelter-in-place orders on March 20, bringing non-essential activities to a partial halt. Furthermore, the use of facemasks in public spaces was made mandatory on April 20. A partial lift of shelter-in-place orders is set to begin on June 1 as worksites adopt social distancing measures to prevent spreading among their customers and employees.

## Model structure

Our model uses 2010 Census data [11] to distribute *N* = 4.3 million inhabitants across 1580 triangular neighborhoods describing the city region [12]. Each neighborhood Ω_*j*_ defines a metapopulation of size *n_j_* and is characterized by demographic and economic variables used to infer the daily mobility habits of its residents (Fig. 1.a). People in each metapopulation are distributed throughout nine epidemiological compartments to keep track of the natural history of COVID-19 (Fig. 1.b, Table S1) such that

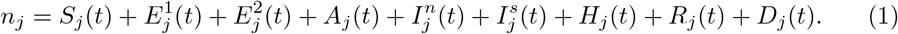

**Figure 1.**
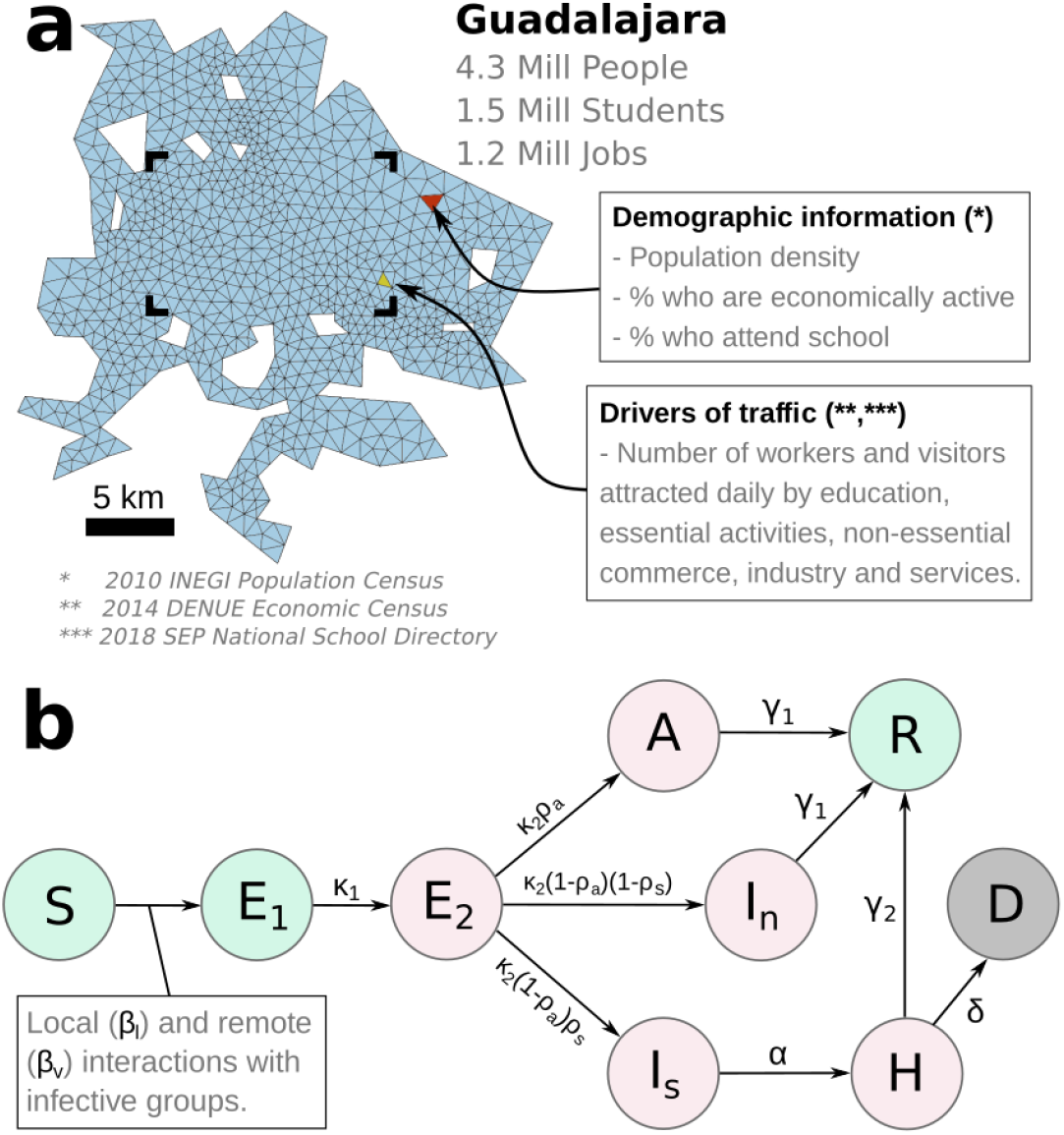
Census and economic data are interpolated onto a triangular grid whose elements represent the different neighborhoods of GDL (panel a). Panel b shows the disease history of COVID-19 as represented in our model; susceptibles (*S_j_*) join 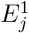 upon exposure and later evolve through infective 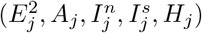 and recovered (*R_j_*) or deceased (*D_j_*) stages. Overall, this approach separates GDL’s 4.3 million inhabitants into 1580 *×* 9 = 14220 subgroups. The rectangular outline in panel a marks the location of data shown in Figure 2.a.

Exposure to SARS-CoV-2 in our model occurs through one of two mechanisms described in Equation (2). The first term 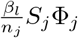 uses a weighted sum Φ_*j*_ (Eq. 6) of the number of residents in Ω_*j*_ within each infective stage of COVID-19 and represents homogeneous mixing within each neighborhood. Thus, the transmission potential *β_l_* controls the frequency of infection within households and between neighbors.

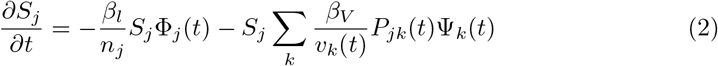

Remote transmission of COVID-19 at places of interest is represented by the second term in the right hand side of Equation 2. Under the formalism of distributed contacts [13,14], this term uses origin-destination matrices *P_jk_*(*t*) to estimate people’s likely travel habits. Given this information, a weighted sum Ψ_*k*_(*t*) (Eq. 7) of the number of infectives visiting locations Ω_*k*_ at time *t* represents the risk of infection for susceptibles traveling to this area. Lastly, the effective number *S_j_P_jk_*(*t*)Ψ_*k*_(*t*) of susceptible-infective pairs at Ω_*k*_ is divided by the total number of visitors *v_k_*(*t*) and multiplied by the transmission potential of remote interactions *β_V_*. To investigate how each term in Equation 2 contributes to epidemic growth, we can calculate their separate contributions to the outbreak’s reproductive number *R_t_* as

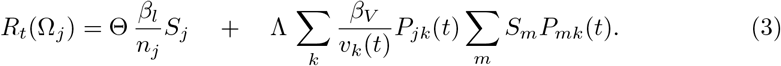

Here, Θ is a coefficient that weighs the relative transmissivity and duration of disease stages in Figure 1.b (Eq. 9). Meanwhile, Λ also incorporates isolation parameters that reduce the frequency with which symptomatic and isolated patients visit public spaces (Eq. 10). Notice that origin-destination matrices in Equations (2),(3) are time-dependent. Because mobility patterns were estimated using records of individual worksites and schools [12], we can use businesses’ NAICS (North American Industrial Classification System) activity codes to represent economic lockdowns targeting specific sectors of the city’s economy (Fig. 2.a).

**Figure 2.**
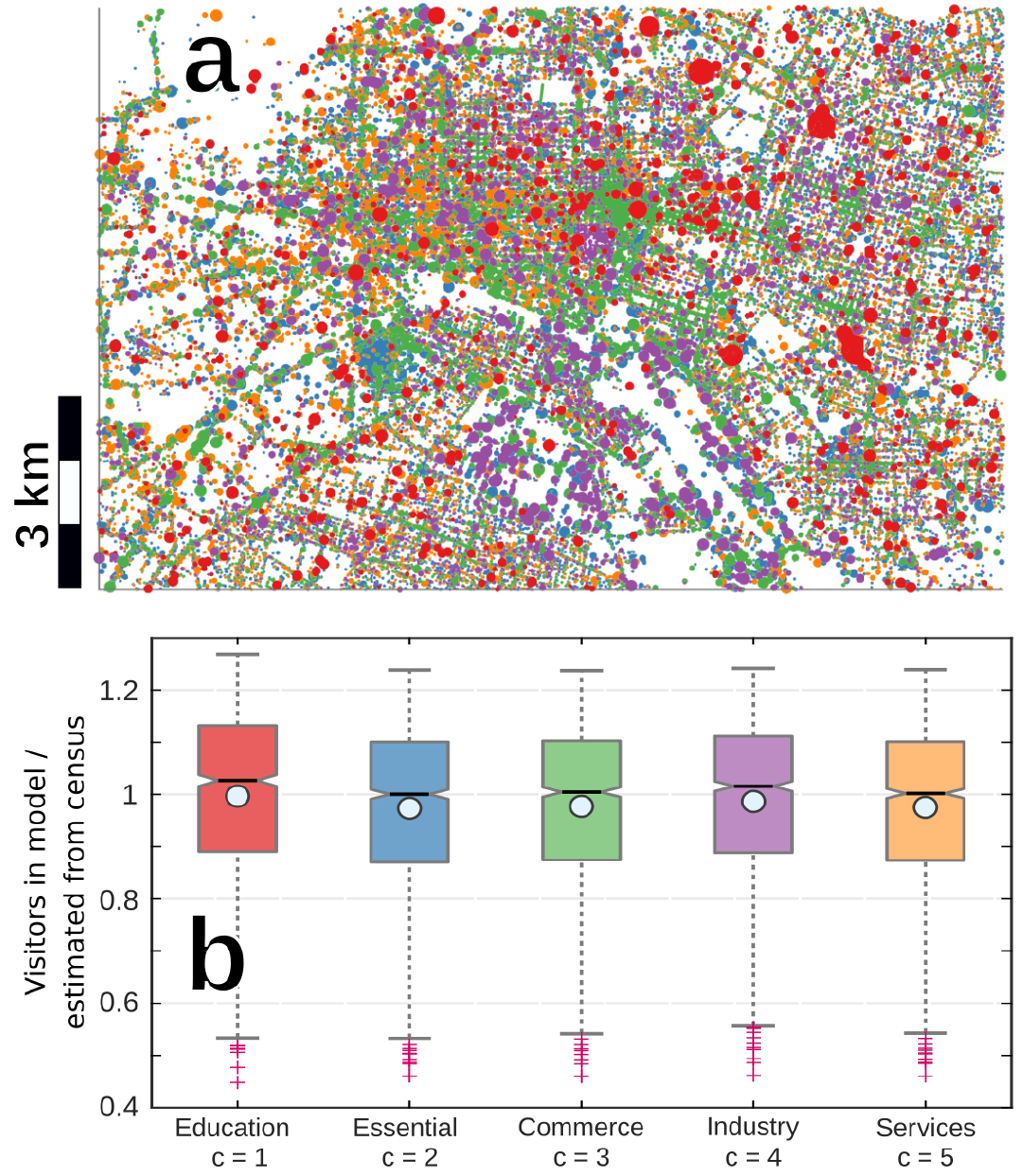
Raw data showing the location, activity sector (color) and number of workers and/or students (circle size) registered by employers and schools that wer used to calculate 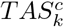 (panel a, see Supplementary Information). The location of this rectangle within the city is shown in Figure 1.a. Box and whisker plots in panel b show quartiles of the ratio 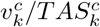 between the model representation 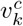 and census estimate 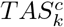 of the number of visitors driven to Ω_*k*_ by establishments in activity sectors *c*. Light circles show that the mean value of this ratio is within 0.05 of 1 for all sectors, indicating that our origin-destination matrices are not biased to overrepresent any of these activity sectors.

## Representing lockdown

Reductions in urban mobility and school closure have been key components for the design of COVID-19 intervention strategies have been noted as a key parameter to quantify the severity of intervention strategies during this outbreak [15–17]. Our model represents changes in the number of daily trips using Equation (4), which uses sector-specific activity levels *a_c_*(*t*) to weigh the contributions 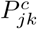 made by each activity sector *c* in the absence of interventions (Table 1, see Supplementary Information).

**Table 1.** Places of interest were classified into five sectors to quantify their separate contributions to urban mobility and epidemic growth. Job data was obtained from the 2014 Economic Census [20], while daily mobility was quantified as the city-wide Trip Attraction Strength 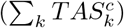 of establishments in each sector.

| Index | Activity sector | Share of job market | Contribution to daily mobility |
| --- | --- | --- | --- |
| $c = 1$ | Education | 8% | 36% |
| $c = 2$ | Essential | 41% | 29% |
| $c = 3$ | Commerce | 19% | 16% |
| $c = 4$ | Services | 19% | 13% |
| $c = 5$ | Industry | 13% | 6% |

Assuming that the city’s baseline status is *a_c_*(*t*) = 1, economic lockdowns can be represented by values *a_c_*(*t*) < 1, thus reducing the magnitude of *P_jk_*(*t*) and subsequent rates of contact in places of interest. Furthermore, because activity sectors are unevenly distributed throughout the city (Fig. 2.a), different values of *a_c_*(*t*) can also transform the topology of contact networks between the city’s neighborhoods.

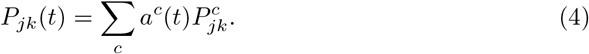

Baseline mobility patterns 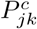 in each sector were inferred using the gravity model in Equation (5). This approach uses the Trip Attraction Strength 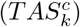 of establishments in each sector to weigh the gravity model of Gonzalez et al. [18]. 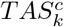was formulated by Jakimavicius and Burinskiene [19] and is an estimate of the daily number of visitors attracted to Ω_*k*_ by activities in sector *c*. It is calculated as a weighted sum of the number of jobs and enrolled students in each neighborhood, which was obtained from the 2014 Economic Census [20] and the 2018 National School Directory [21]. A map of the individual contributions of registered worksites to 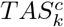 is shown in Figure 2.b.

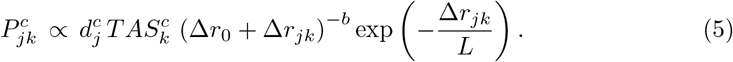

Overall, the gravity model of Equation (5) describes travellers who seek to minimize the distance ∆*r_jk_* between their place of residence Ω_*j*_ and the places they visit Ω_*k*_.

However, they’re willing to travel far given the right incentive (areas with higher 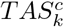). Coefficients 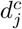 incorporate demographic information that describes the involvement of metapopulations *n_j_* in sector *c*. For example, 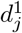 is the fraction of people in Ω_*j*_ who are enrolled in educational programs and was obtained from census data, while 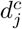, *c* ≠ 1 is a function of the percentage of residents in Ω_*j*_ who are economically active. By using demographic information to refine inferred mobility patterns *P_jk_*(*t*), our framework for disease transmission at places of interest (Eq. 2) can explicitly represent the impacts of economic lockdowns and school closure as they first change mobility patterns.

Mobility patterns used in our model are calibrated so that the number of people 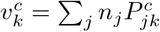 who visit a neighborhood Ω_*k*_ in simulations matches the census estimate 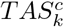 (Fig. 2). Because the mean of ratios 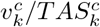 is within 0.05 of 1 for all sectors, we consider that our representation of daily mobility can truthfully represent the relative roles of each activity sector in the city’s transportation network. Notice that activity sectors make contributions to daily mobility 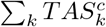 that are disproportionate to number of people that they employ (Table 1). This is caused by secondary visitors like students, customers and suppliers whose presence is necessary to sustain activities in each sector [19]. This implies that business closure in each sector will come at its own economic cost but also yield different public health impacts. For example, the closure of industrial activities in our model is least efficient in our model, as it can affect up to 13% of jobs but only prevent 6% of daily trips.

## Results

Activity levels *a_c_*(*t*) in our simulations were set using Google COVID-19 Mobility reports (www.google.com/covid19/mobility/) for the state of Jalisco between March 5 and May 21 (Fig. 3.a. “Grocery & Pharmacy” data from Google were used to set activity levels in *c* = 2, while “Retail & Recreation” represent *c* = 3 and Google’s “Workplaces” category sets *c* = 4, 5. Mobility scenarios used after May 21 are assumptions based on current policy. Simulations were initialized by the introduction of 1*/*(1 − *ρ_a_*) people into exposed groups for each one of the 66 confirmed cases that were identified as imported from other countries into the State of Jalisco. Imported cases on their dates of arrival to the state of Jalisco, and transmission potentials *β_l_, β_V_* were calibrated to reproduce weekly mortality data and produce a baseline reproduction number 2.5 ≤ *R*_0_ *≤* 3.0 before interventions (Fig. 3). The infection fatality rate (IFR) was set to 0.7%, meaning that 7 out of every 1000 people exposed to SARS-CoV-2 in our simulations die, while the rest recover 1*/κ*_1_ + 1*/κ*_2_ + 1*/γ*_1_ = 12 days after exposure (all model parameters are shown in Table S1). Starting on April 20, we assume that the mandatory use of face covering in public spaces lowers the transmission potential of out-of-home transmission *β_V_* by 50%.

**Figure 3.**
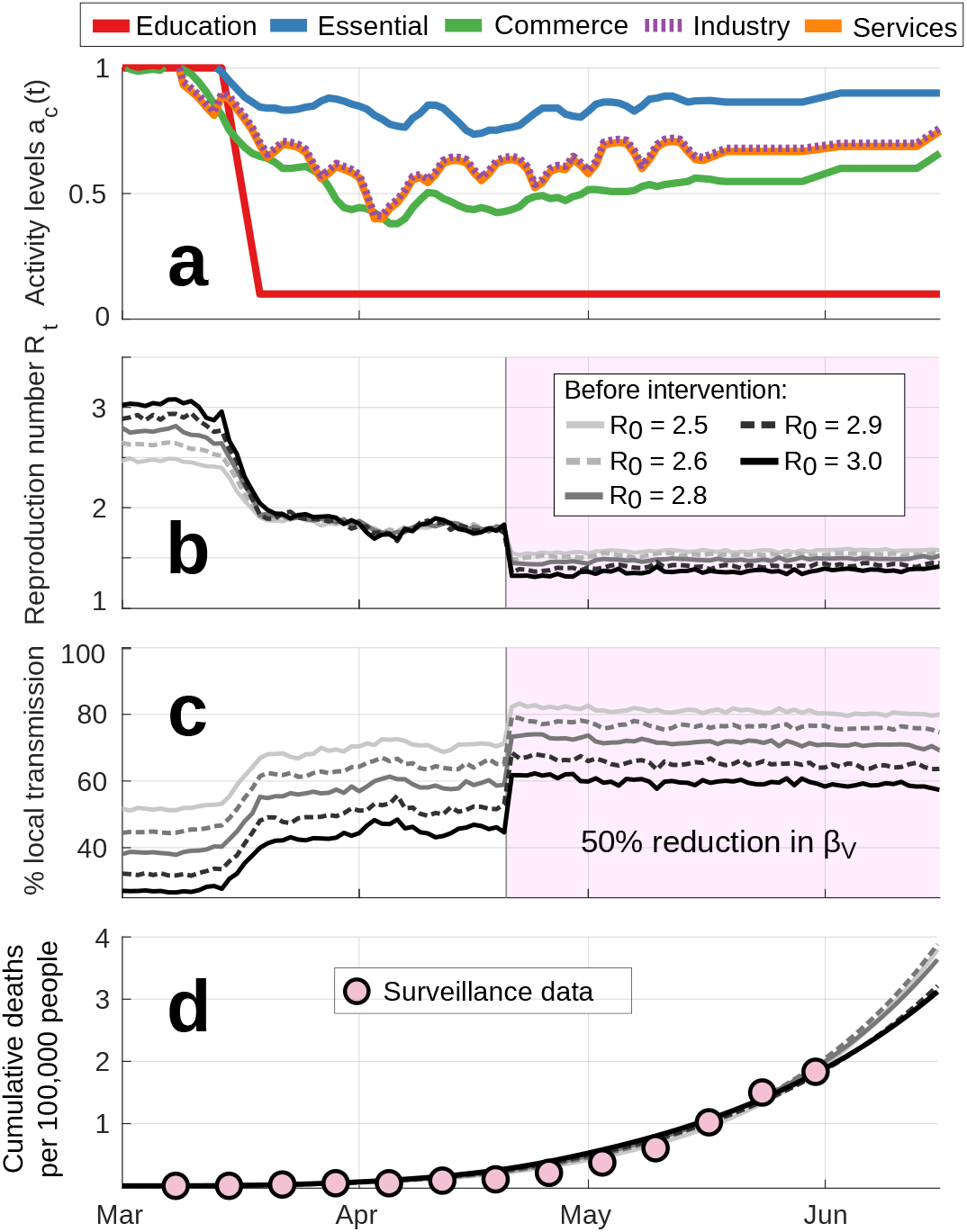
Model simulations of COVID-19 spread in GDL use activity levels *a_c_*(*t*) from Google’s mobility reports (a) to infer temporal changes in the reproduction number *R_t_* (b, Eq. 3). As mobility restrictions become more severe, epidemic growth is increasingly driven by residential transmission (c). Starting on April 20, we represent the mandatory use of face covering in public spaces through a 50% reduction in the corresponding transmission potential *β_V_*. The combination of reduced activity levels *a_c_*(*t*) and a reduced *β_V_* set *R_t_ ∼* 1.4 and suffice to reproduce weekly mortality data in panel **d**.These results suggest that, under current conditions, more than two thirds of COVID-19 transmission occurs within households and between neighbors (**c**).

Notice that increases in the basic reproduction number *R*_0_ must be compensated by changes in the percentage of infections that occur within households and between neighbors (Fig. 3.c). For example, changing *R*_0_ from 2.5 to 2.9 requires that local transmission (*β_l_*) represents 35% (instead of 50%) of epidemic growth in the absence of an intervention. This happens because increasing the value of *β_V_* relative to *β_l_* amplifies the effects of mobility-based interventions *a_c_*(*t*), allowing each simulation to match observed growth rates.

While economic lockdowns and the use of face covering have helped curb the spread of COVID-19 through interactions in GDL’s activity hubs, policy and, social communication have not targeted viral exposure within households. Thus, we assume that *β_l_* has not changed significantly since the beginning of this outbreak and estimate that residential routes of transmission can explain more than 55% of current epidemic growth (Fig. 3.c). We now use *β_V_* = 0.318, *β_l_* = 0.172 (*R*_0_ *∼* 2.9) to demonstrate the potential benefits of public health campaigns that aim to prevent COVID-19 transmission within households.

## Reacting, fast and slow

We now use the disease transmission scheme in Equation (2) to simulate alternate scenarios where mobility-based (*a_c_*(*t*)-based) interventions are combined with aggressive strategies of case isolation (CI). Our model represents improved CI by changing the values of *ρ_s_* and *α*, which were respectively set at *ρ_s_* = 0.1 d^−1^ and *α* = 1*/*3 d^−1^ in our calibration runs (Fig. 3). Campaigns simulated below expand CI to reach 40% or 80% (*ρ_s_* = 0.4, 0.8) of symptomatic patients only 1*/α* = 2 days after symptom onset. While isolated in group *H_j_*, symptomatic patients make reduced contributions to residential (30% of baseline value) and remote (3% of baseline value) disease spreading.

To assess the urgency of CI strategies, we evaluated the effects of deploying such campaigns on June 1, July 1 and August 1. Likewise, and considering that economic lockdowns are bound to be relaxed soon, we tested two alternate scenarios for urban mobility. In the fast scenario (Fig. 4.a,c), gradual reopening of the economy occurs between June 1 and August 1, when economic activities reach their baseline values (Fig. S1.a). The slow scenario (Fig. 4.b,c) extends partial economic lockdowns until September 1 and keeps schools closed through mid-August (Fig. S1.b), while basic education sets *a*_1_(*t*) = 0.6 between June 1 and July 17 in the fast scenario.

**Figure 4.**
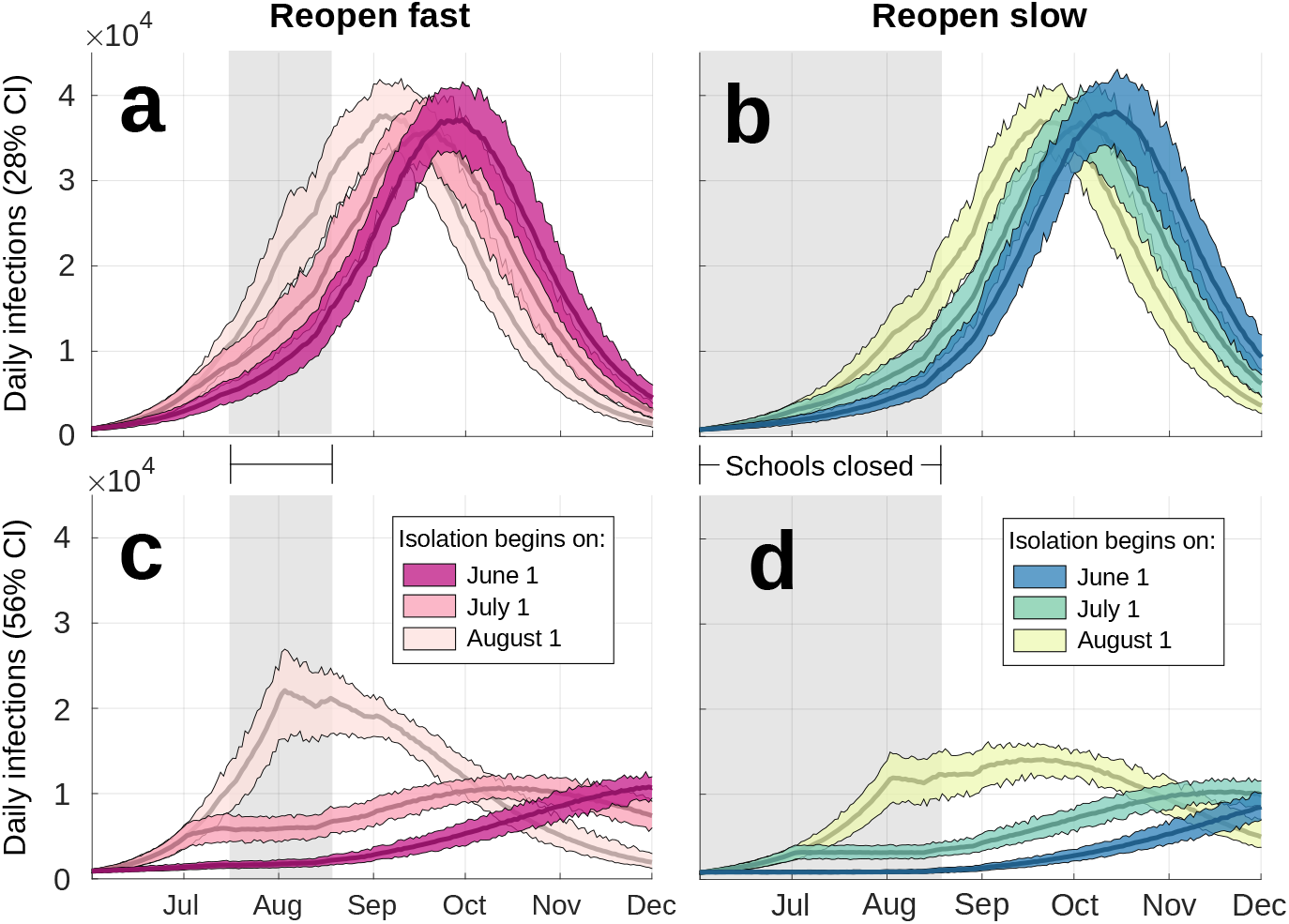
Comparing economic lockdowns and widespread case isolation as COVID-19 mitigation strategies. Differences between the left (a,c) and right (b,d) panels are caused by activity levels *a_c_*(*t*) describing fast and slow paths to reopen the city’s economy (Fig. S1). Simulation ensembles illustrate the effects of improving CI strategies to isolate 28% (a,b) or 56% (c,d) of all people exposed to SARS-CoV-2 on three different dates. Patients are isolated two days after symptoms begin, thus reducing patients’ residential and remote transmission potentials to 10% and 2% their baseline values respectively (*q_H_* = 0.1, *c_H_* = 0.2). Gray shading shows periods when all schools are closed.

Simulation results of daily new cases in Figure 4 suggest that an extended economic lockdown (Figs. 4.b,d) can delay disease spreading after June 1, but will only curb it significantly when combined with widespread CI. Even with 28% CI starting on June 1, the extended lockdown described by activity levels in Figure S1 could sustain more than 30,000 new daily infections for roughly 40 days starting in September (Fig. 4.b). Meanwhile, a fast reopening of economic activities would produce similar infection rates 3 weeks earlier (Fig. 4.a). In contrast, the early adoption of stricter campaigns with 56% CI and extended school closure can produce constant growth (*R_t_ ∼* 1, Figs 4.c,d).

Postponing these measures by only one month could cause a two-fold increase in health care demands throughout June, July and August (Fig. 5).

**Figure 5.**
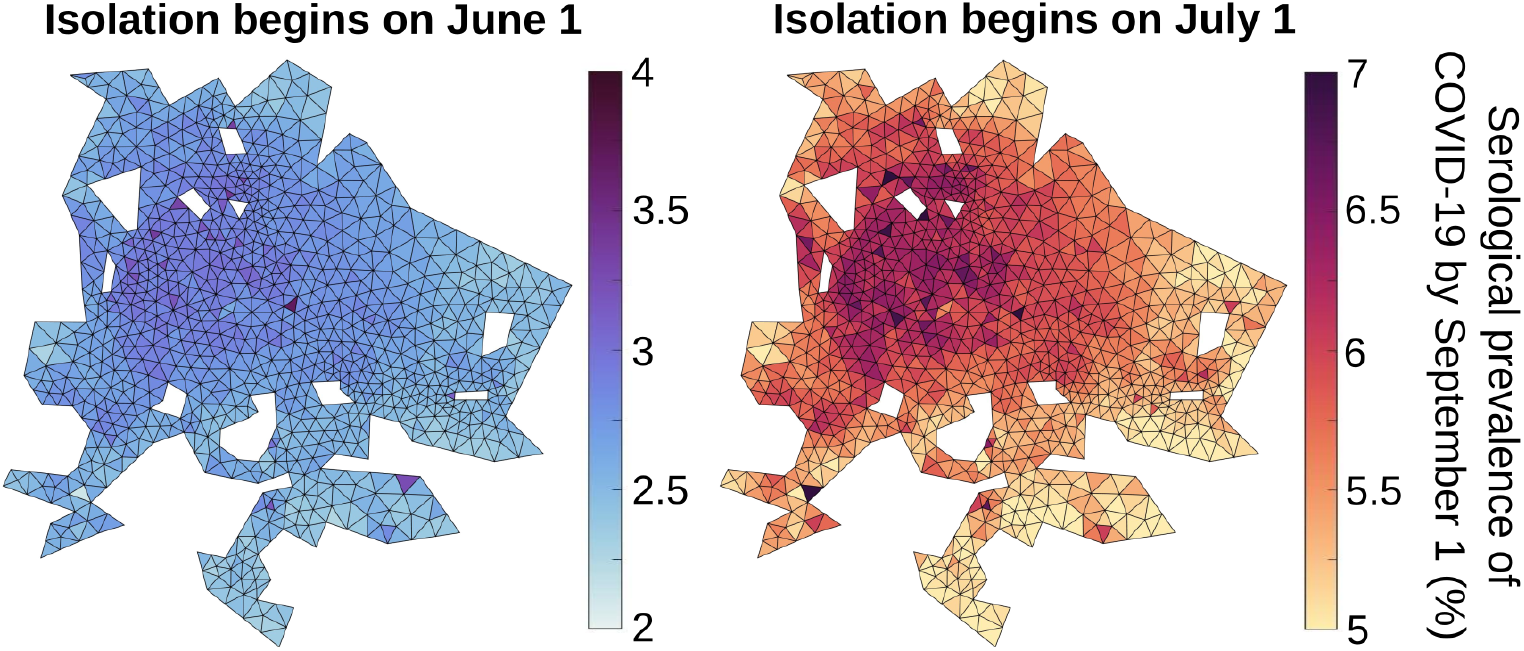
Spatial dependence of the percentage of the population exposed to SARS-CoV-2 by September 1 according to simulations in Figure 4.d. These experiments assume that activity levels follow a slow reopening of the economy (Fig. S1) and that, after improved CI strategies begin, 80% of symptomatic patients (56% CI) are isolated within 2 days of symptom onset.

## Conclusion

We used a high resolution metapopulation model of GDL to represent the spread of COVID-19 under various intervention scenarios (Figs. 3, 4). The disease transmission scheme in Equation (2) represents spreading as the result of both residential (*β_l_*) and remote (*β_V_*) interactions. Non-local spreading follows time-dependent mixing patterns produced by origin-destination matrices inferred from economic and educational records [12] under a distributed-contacts framework [13,14]. By using explicit representations of business and school closures (Eqs. 4, 5, Fig. 2), our simple approach provides realistic estimates of disease spreading under an economic lockdown. Our simulations illustrate that mobility-based interventions are insufficient to stop epidemic growth, as severe COVID-19 outbreaks can be largely fueled by household transmission (Figs. 3.b,c, 6).

**Figure 6.**
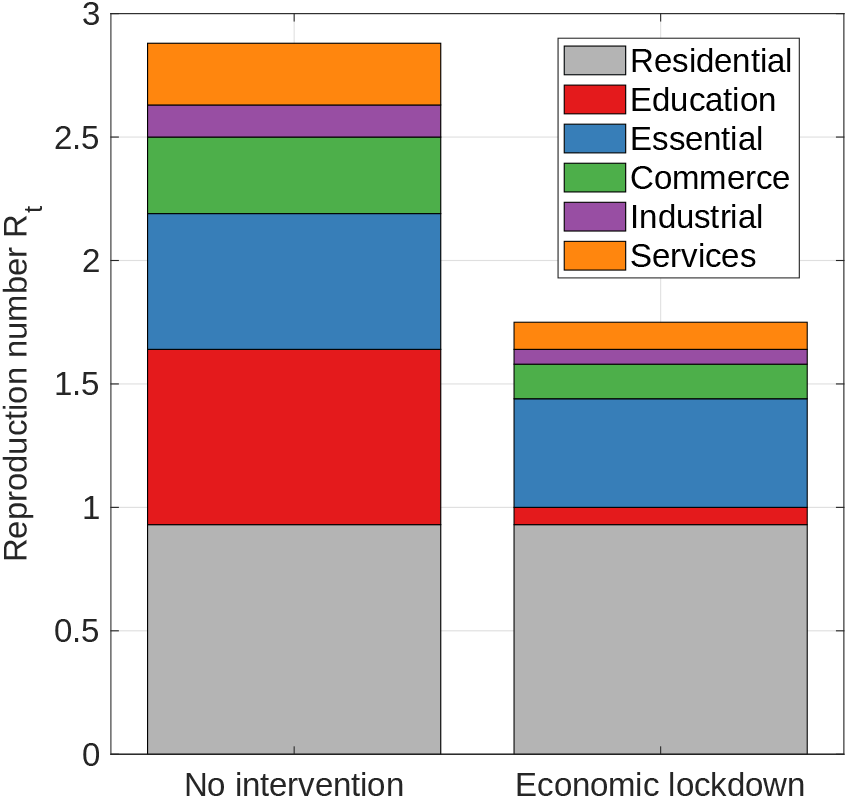
Contributions (see Supplementary Information) to the reproduction number *R_t_* made by residential transmission (*β_l_*) and different economic sectors (*β_V_*, Table 1). Estimates were made using *β_V_* = 0.318 and *β_l_* = 0.172 for a fully-susceptible population in the absence of interventions (*a_c_*(*t*) = 1, left) and during an economic lockdown (*a_c_*(*t*) = [0.1, 0.8, 0.45, 0.45, 0.45], right).

COVID-19 strategies in GDL and countless other cities have focused on reducing daily mobility to suppress disease spreading in public spaces, schools, and workplaces.

All such measures are reflected on the second term of Equation (2) but don’t tackle contagious interactions that occur between household members and other close contacts (first term in Equation 2), which contribute as much as 80% of estimated transmission under lockdown (Fig. 3). This is visualized in Figure 6, which separates the contributions made to *R_t_* by residential transmission and activity sectors with and without an economic lockdown when *β_V_* = 0.318, *β_l_* = 0.172.

By reducing the remote transmission potential *β_V_*, the mandatory use of facemasks in public spaces [22] can further reduce the contribution of remote interactions (Fig.3.b). However, notice that essential activities account for roughly 29% of daily mobility in GDL and cannot be fully suspended during an intervention. This sets an upper bound for the effect of mobility-based strategies that, along with high rates of residential transmission, makes contention unlikely without addressing the mechanisms responsible for disease spreading within households. While lockdowns and social distancing measures as implemented in Wuhan and Shanghai are thought to have stopped the spread of COVID-19 [23], differences in public awareness and compliance, as well as higher rates of case detection and isolation [24] may explain this difference. Wang and coauthors highlight the use of facemasks and frequent disinfection within the households of suspected and confirmed patients [25] as an effective complementary measure to mobility-based interventions.

As financial pressure pushes cities to lift shelter-in-place orders and reopen their economies, we propose the prevention of COVID-19 transmission within residential settings as a necessary complementary measure for successful mitigation. Widespread case isolation (CI) as proposed by many studies [2,3] is unlikely to occur in GDL, as current testing capacity in Mexico is increasingly being reserved for heavily symptomatic patients. Thus, we consider that campaigns must aim to build public awareness about the symptoms of COVID-19 for people to self-diagnose and self-isolate when they’re symptomatic or have interacted with someone who was. Likewise, guidelines for effective self-isolation at home [25] must be heavily publicized and made readily available to everyone, as a majority of cases in developing countries is likely to go undetected and many people still doubt that COVID-19 threatens their own health [26].

We conclude that urgent action is necessary to prevent COVID-19 transmission within households in GDL and other cities within the early stages of this outbreak. As of June 1, we estimate that 0.5-0.9% of the city’s inhabitants have been exposed to SARS-CoV-2, but such low levels of incidence can give the wrong impression that public health strategies used to date are sufficient to contain the epidemic. However, early adoption of widespread CI and other mitigation strategies that prevent disease spreading within residential settings can amplify the effects of mobility-based strategies and social distancing in public spaces (Fig. 5).

The reality of COVID-19 in urban areas is one of drastic inequality in access to healthcare and the ability to self-isolate. Our simulations don’t account for all such heterogeneities, which can largely impact the evolution of an infectious outbreak [27–29]. While our estimates of the reproduction number *R_t_* (Fig. 3.b) are sensible to numerous model parameters and the introduction of imported cases, the conclusion that household transmission is playing an increasing role in epidemic growth (Fig. 4.c) follows from the fact that current policy in GDL only targets spreading in public spaces.

Billions around the world committed to stay home over the past months, and social distancing measures have largely helped slow the spread of COVID-19 in schools, worksites and other places of interest. Nonetheless, as the epidemic progresses and household transmission plays an increasing role in spreading disease, communication campaigns must bring attention to guidelines for the early assessment of symptoms and successful self-isolation at home. This is specially crucial in developing and impoverished countries like Mexico, where diagnosis, treatment and clinical isolation are increasingly unattainable. COVID-19 continues to seed infectious clusters throughout metropolitan areas worldwide, many of which will likely grow into regional-scale outbreaks unless we flatten the curve both inside and outside our homes.

## Data Availability

All data used in this study were retrieved from public repositories and government websites.

https://www.inegi.org.mx/programas/ccpv/2010/

https://www.inegi.org.mx/app/mapa/denue/

https://coronavirus.gob.mx/datos/

https://www.siged.sep.gob.mx/SIGED/escuelas.html

## Supporting information

**Figure s1:**
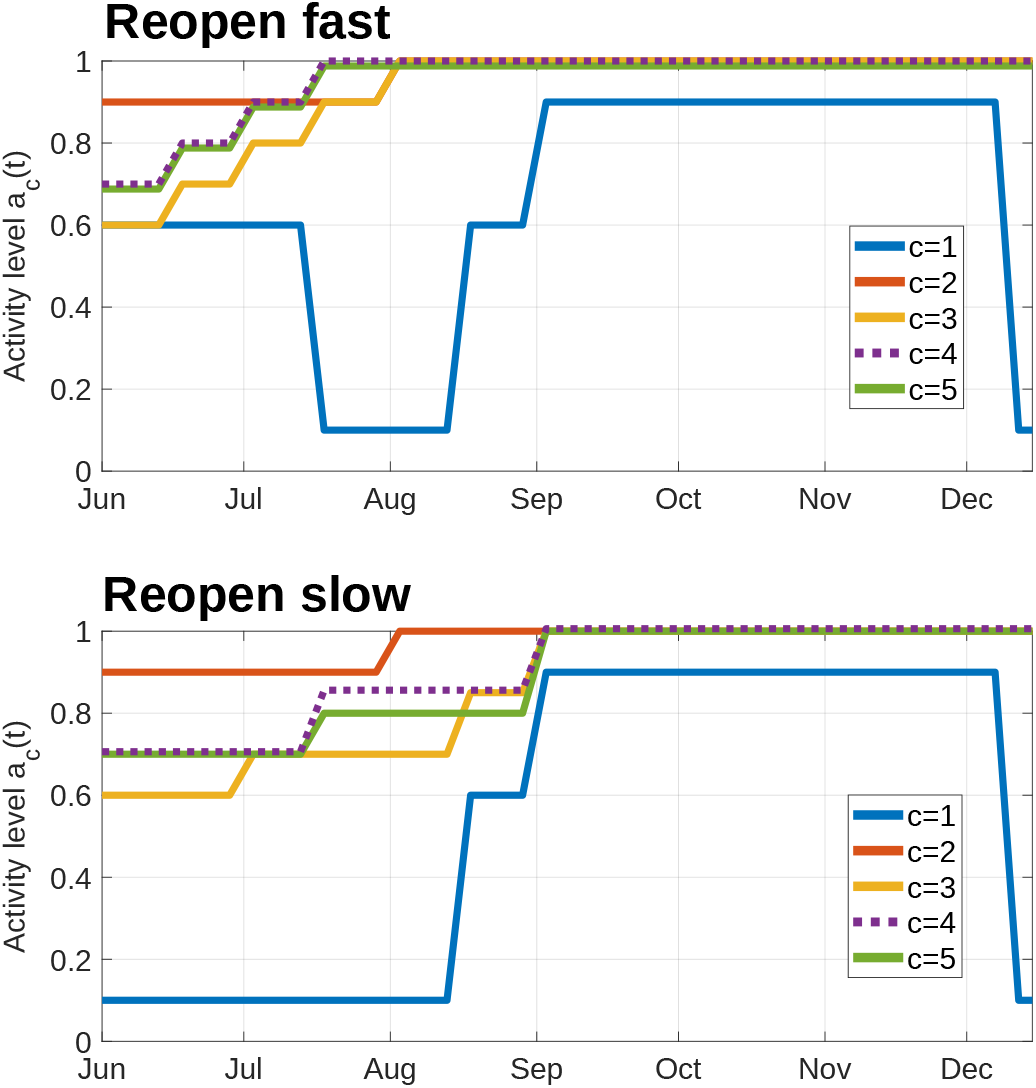
Mobility scenarios to represent fast (upper) and slow (reopening) of the city’s economy. Simulation results under these activity levels *a_c_*(*t*) and different strategies for case isolation are shown in Figure 4, while the full spatial dependence of solutions is visualized in Figure 5

## S1 Appendix

The weighted presence Φ_*j*_(*t*) of infective groups within their own neighborhoods Ω_*j*_ is written as

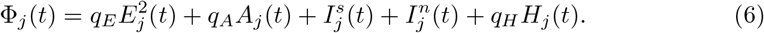

When infectives travel to locations Ω_*k*_, their bulk potential for disease transmission in Equation (2) is given by

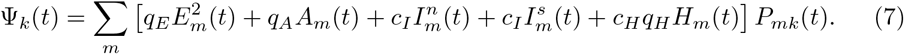

Coefficients *q_E_* = 0.2, *q_A_* = 0.5, *q_H_* = 0.1 account for changes in the transmissibility of COVID-19 throughout different stages of the disease. Likewise, *c_I_* = 0.7, *c_H_* = 0.2 are isolation coefficients that introduce reductions in the relative mobility of individuals when they become symptomatic or self-isolated.

The number of visitors at a destination Ω_*k*_, as used in Equations (2), (3) is given by

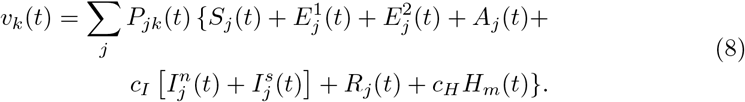

Coefficients Θ (Eq. 9) and Λ (Eq. 10) account for the duration and relative transmissibility of patients within each disease stage, necessary to calculate the reproduction number *R_t_* (Eq. 3).

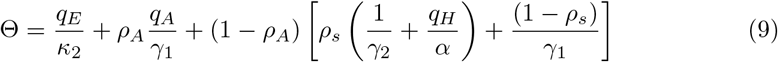

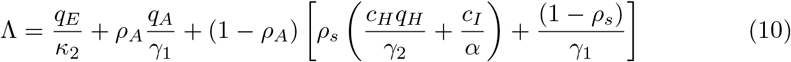

To separate the contributions 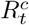 made to the reproduction number *R_t_* by different economic sectors in Figure 6, we substituted infective mobility matrices 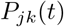 for their sector-specific counterparts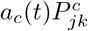 in Equation (3). Origin-destination matrices 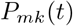 for susceptibles and the form of *v_k_*(*t*) were unchanged.

As defined in previous studies [12,19], the Trip Attraction Strength (*T AS_k_*) of a neighborhood Ω_*k*_ is calculated as the weighted sum

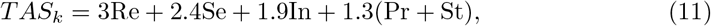

where parameters Re, Se, In and Pr denote the number of jobs registered at Ω_*k*_ by retail, service, industrial and primary activity organizations respectively. Similarly, St is the number of students enrolled at educational institutions inside the same area. We used georeferenced employer records from the 2014 Economic Census [20] and the 2018 National Directory of Schools [21] to allocate workers and students throughout the city’s neighborhoods. NAICS (North American Industrial Classification System) activity codes were used to assign employers across mobility categories in Equation (11) and Table (1). Raw data used to calculate *T AS_k_* are shown separated by activity sector are shown in Figure 2.

**Table S1:** Model parameters. Model parameters used to describe the disease history of COVID-19 as represented in Figure 1.b.

| Model parameter | Description | Value |
| --- | --- | --- |
| $1/\kappa_1$ | Average length of latent period | 2.5 days |
| $1/\kappa_2$ | Average length of infectiousness prior to symptom onset | 2.5 days |
| $\rho_a$ | Proportion of exposed individuals who become asymptotically infected | 0.3 |
| $\rho_s$ | Proportion of fully infectious individuals who will be screened and isolated | 0.1 |
| $1/\alpha$ | Average time from symptom onset to diagnosis | 3 days |
| $1/\gamma_1$ | Average time from illness onset to recovery | 7 days |
| $1/\gamma_2$ | Average length of time from diagnosis to recovery | 4 days |
| $\delta$ | Disease-induced death rate within hospitals | $\text{IFR} \frac{\gamma_2}{\rho_s(1-\rho_a)}$ |
| IFR | Infection fatality rate | 0.7% |
| $q_E, q_A, q_H$ | Relative transmissibility of exposed (E), asymptomatic (A), and isolated (H) individuals. | 0.2, 0.5, 0.1 |
| $c_I, c_H$ | Relative reductions in the mobility of fully infectious (I) and isolated (H) individuals | 0.7, 0.2 |

## Acknowledgments

We thank the faculty and students at Universidad de Guadalajara who volunteer without pay to collect and process diagnostic samples for SARS-CoV-2. The authors declare no conflict of interest.

